# Development of a Communication Characteristics Module for Autistic Adults and the Creation of a *Communication Profile* Variable

**DOI:** 10.64898/2026.10.15.26365687

**Authors:** Rachel Kripke-Ludwig, Joseph Vera, Ian Moura, Steven K. Kapp, Zack Siddeek, Joelle Maslak, Anna Furra Wallington, Shannon Des Roches Rosa, Clarissa C. Kripke, Christina Nicolaidis

## Abstract

**Purpose:** Autistic adults have diverse communication characteristics that are not well captured by standard demographic measures. Communication affects self-determination, relationships, access to healthcare, and services. Characterizing communication differences may improve research and intervention targeting for meaningful subgroups of autistic adults.

**Methods:** The Academic Autism Spectrum Partnership in Research and Education (AASPIRE) Outcomes Project team used a community based participatory research (CBPR) approach to develop a self- or caregiver-report module to characterize communication among autistic adults. The 10-item module asks about receptive and expressive communication, literacy, speech reliability, usage of digital communication, use of alternatives to speech, and communication fluency. It can be completed directly, with or without support, or via a caregiver. We used baseline survey data from 844 autistic adults to evaluate the module and derived a five-level variable, *Communication Profile*, from selected module items to stratify participants. We examined completion feasibility, response consistency, and construct validity. We also assessed test-retest reliability in a 15% random subsample who completed a 2-week retest following the 6-month survey, with oversampling of caregiver-report respondents.

**Results:** The Communication Characteristics module demonstrated high completion feasibility, minimal systematic error, and strong construct validity. The *Communication Profile* variable stratified participants into five meaningful communication profiles and demonstrated strong construct validity and substantial test-retest reliability (weighted *K =* .69).

**Conclusion:** The Communication Characteristics module and *Communication Profile* variable provide a valid approach for characterizing communication diversity among autistic adults for research, clinical, and service settings. The full AASPIRE Measurement Toolkit is available at https://aaspire.org/measurement.

**Lay Summary:** Autistic people communicate in many different ways, and a person’s communication may change in different situations. We developed and tested a set of survey questions to describe an individual’s communication. We also used the results to describe 5 groups of autistic people based on how they communicate, without treating these groups as different types of autism.

---

Autism is characterized by substantial heterogeneity in neurological, biological, and genetic traits that influence social communication and interaction (Waterhouse et al., 2016; Waterhouse, 2022). Differences in sensory processing, motor coordination, cognition, and perception can interact in complex ways to shape communicative behavior (Ahtam et al., 2020; Kadlaskar et al., 2023; Scheerer et al., 2021; Torres et al., 2013; Torres et al., 2016). This heterogeneity contributes to wide variation in how autistic individuals express and receive information and, in the supports, and accommodations that facilitate successful communication (Brignell, 2018; Cummins et al., 2020; Jaswal et al., 2020; Marocchini & Baldin, 2024; Pirinen et al., 2023; Pope et al., 2025; Schuck et al., 2025; Wasiuk et al., 2025).

Communication differences among autistic people manifest across expressive and receptive language, pragmatic language use, speech production, and multimodal communication (de Marchena et al., 2025; Pickles et al., 2014). Some autistic individuals speak fluently but experience difficulties with pragmatic language, conversational reciprocity, or interpreting nonverbal cues. Others communicate primarily through augmentative and alternative communication (AAC), writing, or other non-speech modalities (Beukelman & Light, 2020). Still others demonstrate context-dependent communication abilities, such as speaking fluently in structured environments while experiencing substantial communication barriers in everyday settings. Speech may be “expensive”, requiring substantial effort; “intermittent or unreliable”, varying across contexts or over time; or, “insufficient” to fully communicate a person’s thoughts, needs, or experiences (Zisk & Dalton, 2019). These experiences may be shaped by hearing or auditory processing differences, fatigue, sensory overload, emotional dysregulation, or challenges with communication partners who do not accommodate different communication modalities (Ruiz et al., 2023). Communication differences for autistic people may also co-occur with intellectual disability, sensory challenges, or motor differences that affect expressive or receptive language abilities (Craig et al., 2021; Jaswal et al., 2020; Jaswal et al., 2024).

Because communication abilities vary widely, the interventions, accommodations, supports, and professional training needed to improve health and social outcomes may also differ substantially. However, research and service systems often lack meaningful ways to characterize communication differences within autistic populations, particularly among adults (Morrison et al., 2019). Improving outcomes for autistic adults therefore requires approaches that can characterize communication differences beyond broad classifications of autism or speech status.

## Complexity of Categorizing Communication Characteristics

Communication is inherently multidimensional and bidirectional. It includes receptive and expressive processes and multiple linguistic domains such as syntax, semantics, and pragmatics. Effective communication also often depends on prosodic, paralinguistic, and nonverbal features such as tone, pitch, rhythm, facial expression, and gesture (Holbrook & Israelsen, 2020; Zhang et al., 2022; Trayvick et al., 2024). And to communicate effectively, most individuals communicate using multiple modalities. Spoken language may be combined with facial expressions, vocalizations, gestures, writing, or AAC, and these modalities may vary by context or communication partner (Beukelman & Light, 2020). Thus, communication abilities may fluctuate across time, environmental demands, and available human or technological support.

Together, these factors make it difficult to describe communication using a single dimension, such as speech production. Evaluating interventions, accommodations, and services requires meaningful ways to identify communication profiles within the autistic population. Interventions that benefit one group of communicators may be ineffective or inaccessible for others, and studies that treat autistic participants as homogeneous communicators may obscure important differences in communication experiences and outcomes.

## Previous Attempts to Categorize Autistic Communicators

Several efforts have attempted to characterize communication differences within autistic populations, although important limitations remain. In a systematic review of patient-reported outcome measures (PROMs) used with adults with communication disabilities, Van Ewijk et al. (2025) found that none demonstrated adequate content validity and called for participatory approaches involving disabled communities. Girolamo et al. (2024) similarly found inconsistent reporting of structural language characteristics in autism.

Others have tried to develop frameworks for communication differences in autistic children. Tajik-Parvinchi et al. (2023) validated the Autism Classification System of Functioning: Social Communication for pediatric populations, which categorizes communication across five levels while acknowledging contextual variability and multimodal communication. However, it was not designed for adults and relies on caregiver reports of communication behaviors across modalities. Other empirical approaches have used clustering to subgroup autistic children based on language and speech characteristics. Broome et al. (2021), for example, identified three clusters in a small cohort: high language and speech ability; low expressive language with relatively stronger receptive language; and broadly low language and speech development. Such approaches are difficult to apply to people with substantial expressive and motor communication challenges because receptive abilities may be difficult to demonstrate. Further, many non-speaking autistic people have difficulty with speech and motor praxis and therefore may be unable to demonstrate what they understand (Jaswal et al., 2026).

Theoretical proposals have also described communication profiles in autism. Vogindroukas et al. (2022) proposed four profiles: pragmatic language difficulties without structural language impairment; autism with co-occurring language disorders; autism with intellectual disability; and limited or absent use of spoken language, including individuals with speech praxis disorders. However, theoretical proposals cannot be adequately validated when communication and cognitive assessment tools themselves may not accurately assess people with substantial verbal and motor praxis difficulties (Bagnato & Neisworth, 1994; Courchesne et al., 2015; Ellert et al., 2026).

Many approaches to subgrouping autistic individuals also rely on broad categories based on support needs or functional impairment (American Psychiatric Association, 2022; Lord et al., 2022; Interagency Autism Coordinating Committee, 2026). However, these classifications risk treating support needs or functional impairment as proxies for overall ability, potentially obscuring important variation in how autistic adults communicate and reinforcing assumptions of reduced competence among those with greater support needs. There is therefore a need for a measure focused specifically on communication characteristics that can be considered separately from other traits, characteristics, and co-occurring conditions when examining life outcomes.

## The Present Study

To address this gap, the present study describes the development and assessment of the AASPIRE Communication Characteristics module as part of the larger AASPIRE Outcomes Project. The AASPIRE Outcomes Project used a Community-Based Participatory Research (CBPR) approach to develop and test the AASPIRE Measurement Toolkit (www.aaspire.org/measurement), a set of PROMs and demographic modules for evaluating services for autistic adults. Methods and results of the overall project are described elsewhere (Nicolaidis et al., 2025; Nicolaidis et al., 2026a; Nicolaidis et al., 2026b).

Our goals for this study were to 1) evaluate the full Communication Characteristics demographic module and 2) propose a single variable to stratify autistic adults by communication profile. Importantly, our proposed framework does not rely solely on speech production as a proxy for overall communication ability. The ability to produce speech does not necessarily reflect the ability to express complex thoughts across contexts or modalities, and speech may not be an individual’s most robust or accessible form of communication.

## Methods

### Study Design and Setting

This study evaluated the psychometric characteristics of the Communication Characteristics module; a demographic instrument developed as part of the AASPIRE Outcomes Project (Nicolaidis, 2026a). The module was designed to describe heterogeneity in communication among autistic adults across receptive language, expressive language, literacy (i.e., reading and writing), and alternative communication modalities. Although originally developed to support subgroup characterization within the broader project, this study examined the module as a standalone instrument and assessed the completion feasibility, reliability, and construct validity of both the module and a derived stratification variable, *Communication Profile*, created from module responses.

The Institutional Review Board at Portland State University approved the study. Other participating institutions ceded oversight to Portland State University, and the study conformed to the U.S. Federal Policy for the Protection of Human Subjects. Participants or their legally authorized representatives provided informed consent online, by telephone, or in person. Details about the accessible informed consent and authorization process (Beck et al., 2025) and data sharing process (Grant et al., 2026) are available elsewhere.

### Community-Based Participatory Research Approach

The AASPIRE partnership is an established research collaboration with longstanding procedures for conducting CBPR with autistic adults. The partnership’s inclusive research procedures are described elsewhere (Nicolaidis et al., 2019). The AASPIRE Outcomes Project team included autistic and non-autistic academic partners, students and staff, autistic community partners, parents of autistic adults with high communication support needs, clinicians, and disability service providers. Autistic community partners for the Outcomes Project included 10 autistic adults with varying communication profiles, including a minimally speaking autistic adult, adults with unreliable or “expensive” speech, full- and part-time AAC users, people with auditory processing differences, and people with different reading, writing, and support needs.

The full AASPIRE Outcomes Project team created the Communication Characteristics module by adapting and expanding communication-related items from the AASPIRE Autism Healthcare Accommodations Tool (Nicolaidis et al., 2016). A smaller workgroup, including the principal investigator, a speech therapist, a minimally speaking autistic adult, supporters of autistic adults, and physicians who care for autistic adults, subsequently developed and derived the demographic variable. The *Communication Profile* variable used responses from selected module questions. All team members reached consensus on the module items and approved the derived variable, and all had the opportunity to be co-authors on this manuscript.

### Participants and Procedures

Participants were autistic adults enrolled in the AASPIRE Outcomes Project who completed the Communication Characteristics module during at least the baseline study wave (*n* = 844). Most participants completed surveys without support (*n* = 588). If needed, they could get help from their own supporters or a research assistant (*n* = 187). In cases (*n* = 69) where a person would not be able to complete the survey themselves even with support, caregivers used a caregiver report version to answer on behalf of the autistic person. The caregiver report survey was similar to the direct self-report version, but phrased questions in the third person. Recruitment occurred across two healthcare systems, two disability service systems, and within the broader autistic community across the United States. Details about recruitment and cohort creation are described elsewhere (Nicolaidis et al., 2026a). The larger cohort completed surveys at three, primary timepoints over 12-18 months. A 15% random sample, oversampled for caregiver-report respondents, repeated the survey 2-weeks after the 2nd time point to assess test-retest reliability. We used baseline data to evaluate the module and develop the *Communication Profile* variable; we used test-retest data to evaluate stability.

### Communication Characteristics Module Development

The Communication Characteristics module is a free, 10-question demographic instrument designed to characterize communication among autistic adults (Supplement A). Because communication dimensions cannot be represented by a single construct, the module captures several related but distinct aspects relevant to daily life.

The first four questions ask participants to describe how they understand and use spoken language in good conditions and typical daily life. These questions use four-point ordinal response options ranging from no difficulty to very limited understanding or severe difficulty speaking. Questions five and six assess reading and writing skills in good conditions using four-point ordinal scales. Question seven asks how often speech is a reliable form of communication using a five-point Likert scale. Question eight is conditionally administered when the response to question 7 states that speech is not reliable “all or almost all of the time” and it asks about alternatives to speech, including AAC. Question nine is a check-all-that-apply item assessing reliable digital communication modalities, including telephone, videoconferencing, text messaging, and email. Question ten asks participants to describe their overall communication ability in their preferred communication modality under good conditions using a four-category ordinal response scale.

Rather than focus on traditional clinician-defined and typically deficit-focused descriptors such as vocabulary size, utterance length, or articulation accuracy, our module emphasizes communication characteristics identified as meaningful by autistic adults, including the ability to understand spoken speech and express thoughts across modalities.

### *Communication Profile* Variable Development

The smaller AASPIRE workgroup created the *Communication Profile* variable as a derived stratification measure to summarize communication characteristics for subgroup analyses within the AASPIRE Outcomes Project and other research using the module. We constructed our variable using an algorithm (Figure 1) based on six selected module items. Because communication abilities are multidimensional and may vary independently, the workgroup did not seek to create a continuous score. Instead, the goal was to generate conceptually interpretable strata for analytic subgrouping.

**Figure 1.**
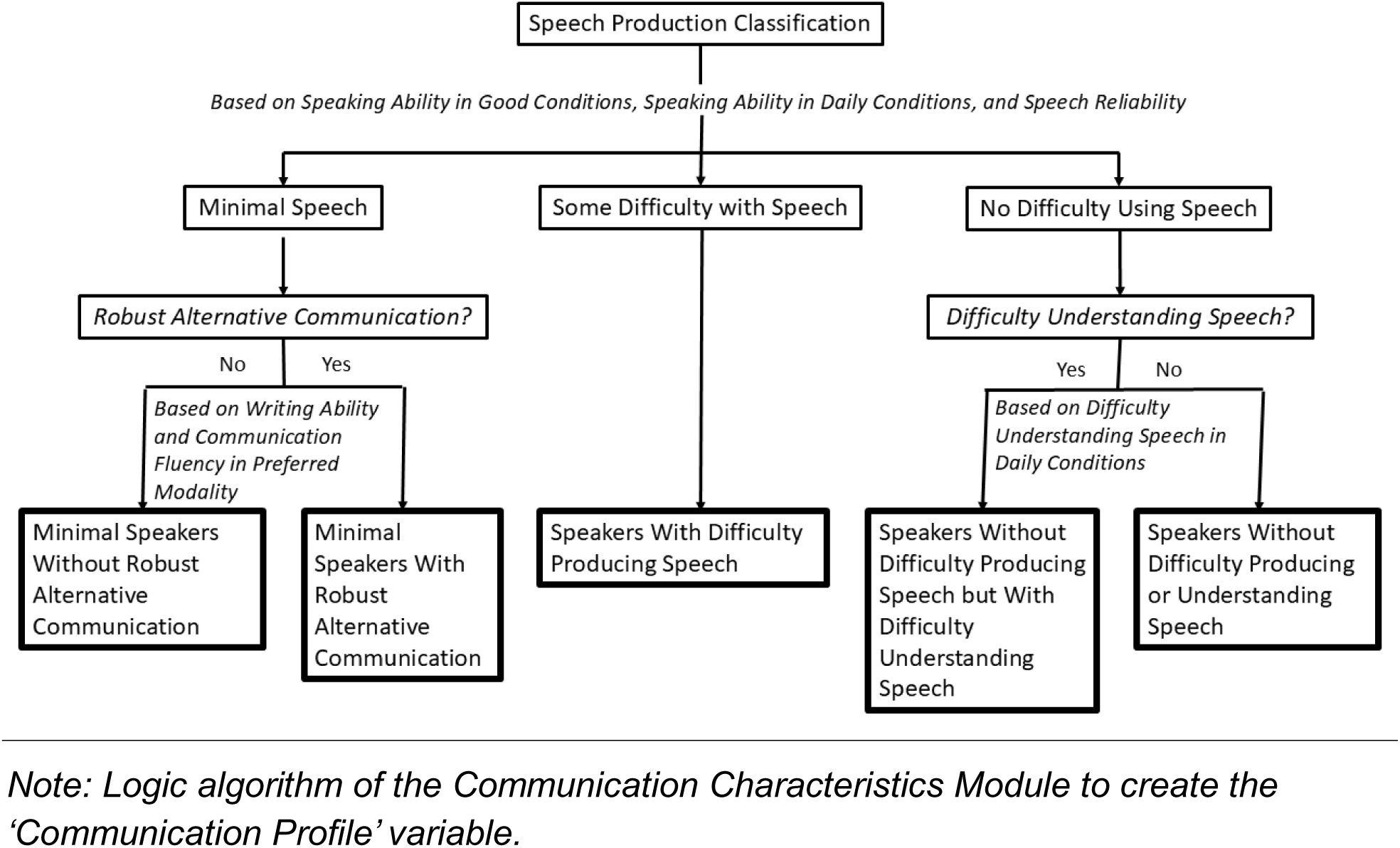
Logic Model to Create the ‘Communication Profile’ Variable.

Our algorithm first categorized participants into three broad levels based on response patterns across items assessing speaking ability in good conditions, speaking ability in daily conditions, and speech reliability: people with minimal speech, people with difficulty using speech, and people without difficulty using speech.

Participants classified as having minimal speech were further divided based on whether they demonstrated a robust alternative communication system, using items assessing writing ability and overall communication fluency in their preferred modality. Participants classified as having no difficulty with speech were also further stratified, according to whether they reported difficulty understanding speech in regular daily conditions, allowing identification of individuals without difficulty producing speech but with auditory processing challenges.

This process resulted in five, mutually-exclusive categories: Minimal Speakers without robust alternative communication; Minimal Speakers with robust alternative communication; Speakers with difficulty producing speech; Speakers without difficulty producing but with difficulty understanding speech; and Speakers without difficulty producing or understanding speech.

### Psychometric Evaluation of the Communication Characteristics Module Items and *Communication Profile* Variable

Because the Communication Characteristics module is a multidomain demographic characterization instrument rather than a unidimensional PROM, we selected psychometric analyses appropriate for descriptive measures with heterogeneous item formats. We used the COSMIN taxonomy to guide evaluation of reliability, completion feasibility, and construct validity (Mokkink et al., 2010). We conducted all analyses using Stata (Version 19) statistical software. Categorical variables were summarized using frequencies and percentages, and *p*-values < .05 were considered statistically significant.

#### Completion Feasibility and Missing Data

We assessed completion feasibility through item-level completion. For each item, we calculated the number of eligible respondents, completed responses, and percentages completed and missing. For conditionally administered items, denominators were limited to participants eligible under the skip logic.

For items with 1 to 5% missingness, we conducted brief subgroup checks to identify systematic nonresponse. For items with greater than 5% missingness, we created binary missingness indicators and used Pearson chi-square tests to examine missingness by survey wave, respondent version, and *Communication Profile*.

#### Qualitative Review of Alternative Communication Write-In Responses

Question eight included an open-ended “Other” response option. We reviewed write-in responses to assess whether existing categories adequately represented alternative communication supports and to identify differences between direct-report and caregiver-report responses. We summarized communication supports or strategies not represented by existing options and compared patterns across respondent versions.

#### Response Consistency and Measurement Error

We assessed logically inconsistent response patterns across paired items addressing communication in good conditions versus regular daily life. Specifically, we examined the proportion of participants who reported better understanding or speaking ability in regular daily life than in good conditions and assessed whether such responses appeared systematic or random.

#### Content Validity

We ensured content validity for the module and the *Communication Profile* variable through the CBPR development process, in which autistic partners identified communication domains relevant to describing their communication and worked as part of the research team to develop and refine item wording.

#### Construct Validity

Because the module is not intended to function as a unidimensional scale, we did not assess internal consistency or factor structure. Instead, we evaluated construct-related validity through *a priori* hypothesis testing. We tested expected associations between related, item pairs using Pearson chi-square tests, including receptive language and literacy, expressive communication and overall communication fluency, and matched items assessing communication in good conditions versus daily life, ability to read versus ability to write, and frequency speech is reliable versus communication fluency.

We assessed known-groups validity by comparing item-response distributions across respondent versions (i.e., direct self-report, direct report with support, and caregiver report). We hypothesized that caregiver-report respondents would more often represent autistic adults with greater communication support needs and therefore endorse greater communication challenges.

To test validity of the *Communication Profile* variable, we used *a priori* hypotheses regarding associations between *Communication Profile* and respondent version, reading ability, intellectual disability status, understanding spoken language, support needs, healthcare communication barriers, and overall barriers to communication. Depending on variable type, we used Pearson chi-square tests or independent-samples *t*-tests. For significant comparisons, we calculated Cramér’s *V*.

#### Test-Retest Reliability

We assessed test-retest reliability by comparing individual items and *Communication Profile* classifications from the Time 2 survey and the 2-week retest administration using Cohen’s kappa coefficients. We calculated linear weighted kappas to evaluate whether disagreements occurred between adjacent or more distant profile categories.

## Results

### Sample Characteristics

A total of 870 autistic adults participated in the AASPIRE Outcomes Toolkit Study (Nicolaidis et al., 2026a). Of these, 844 participants (97.0%) completed the Communication Characteristics module and were included in analyses. The sample demonstrated substantial communication heterogeneity, although minimally speaking participants were somewhat underrepresented relative to estimates of the autistic population (de Marchena et al., 2025). Table 1 shows the overall demographics of our sample.

**Table 1.**
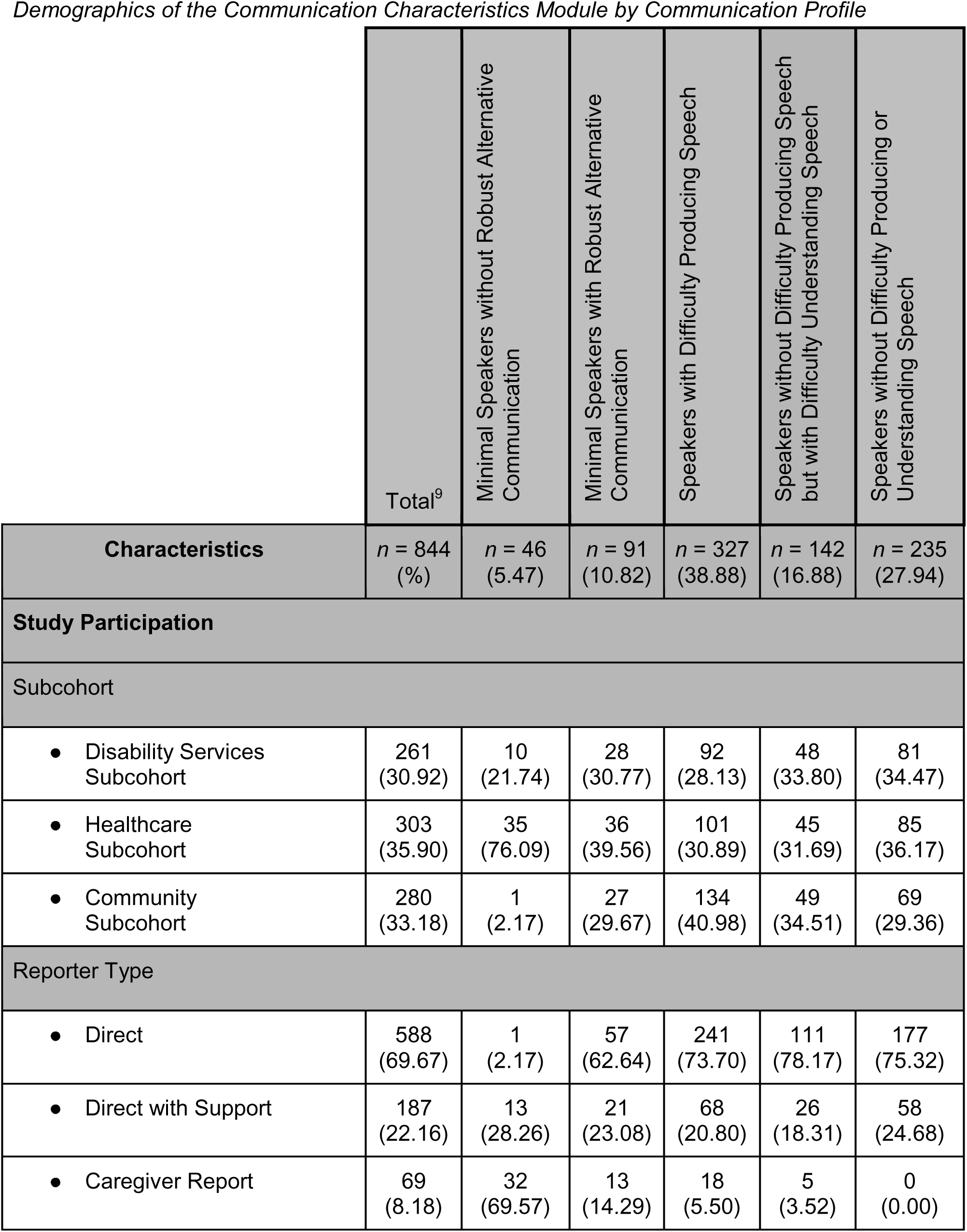
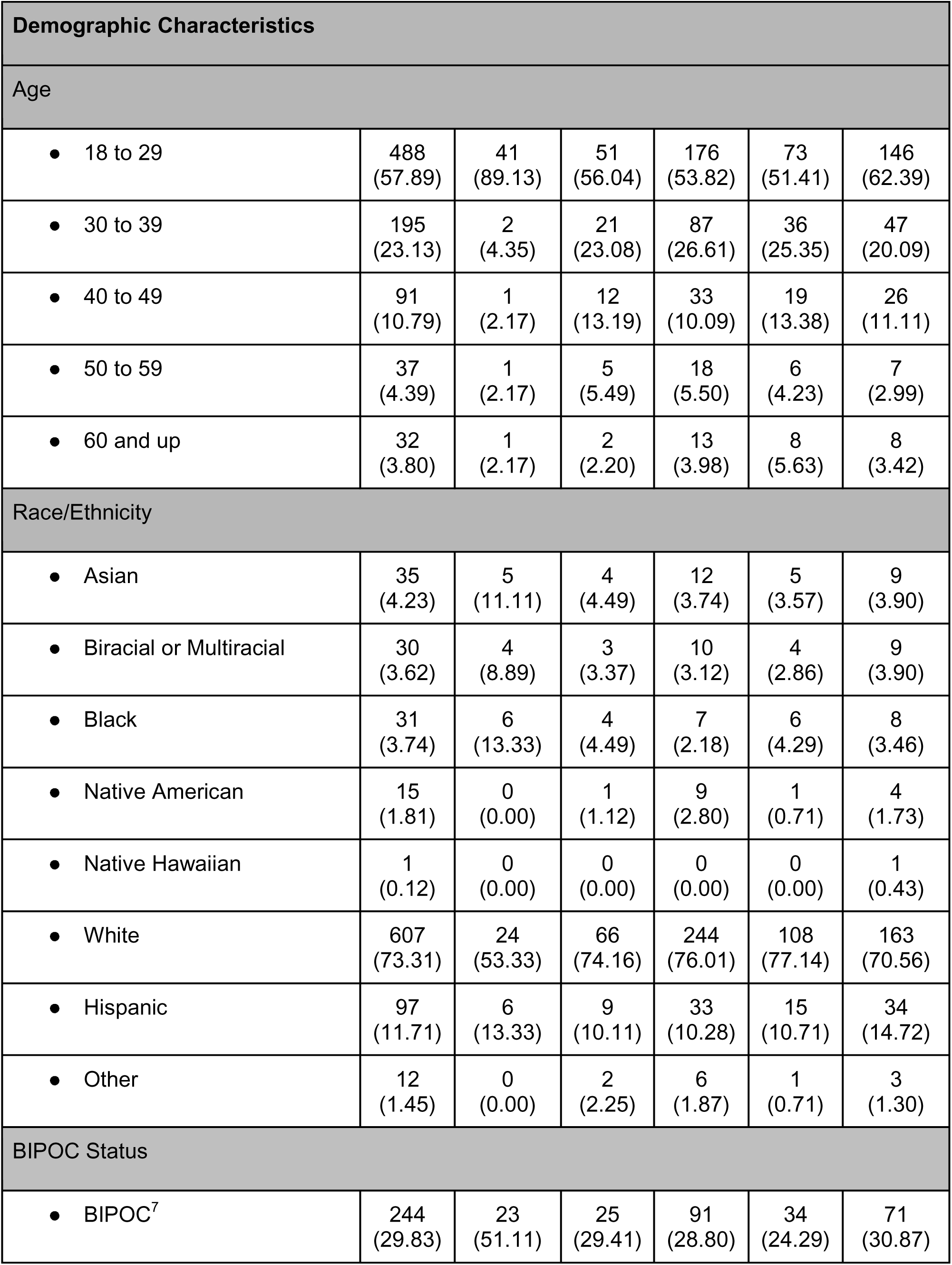
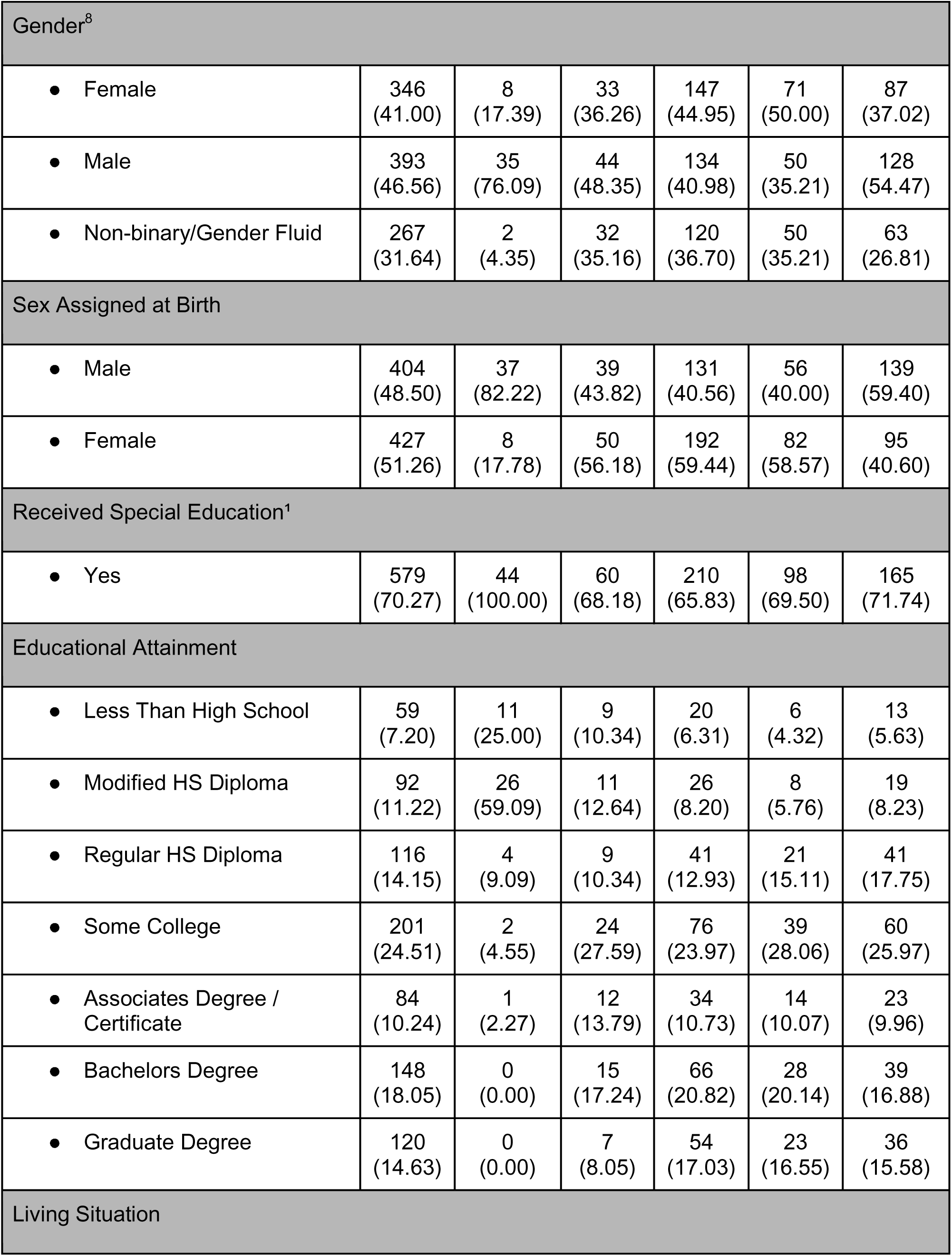
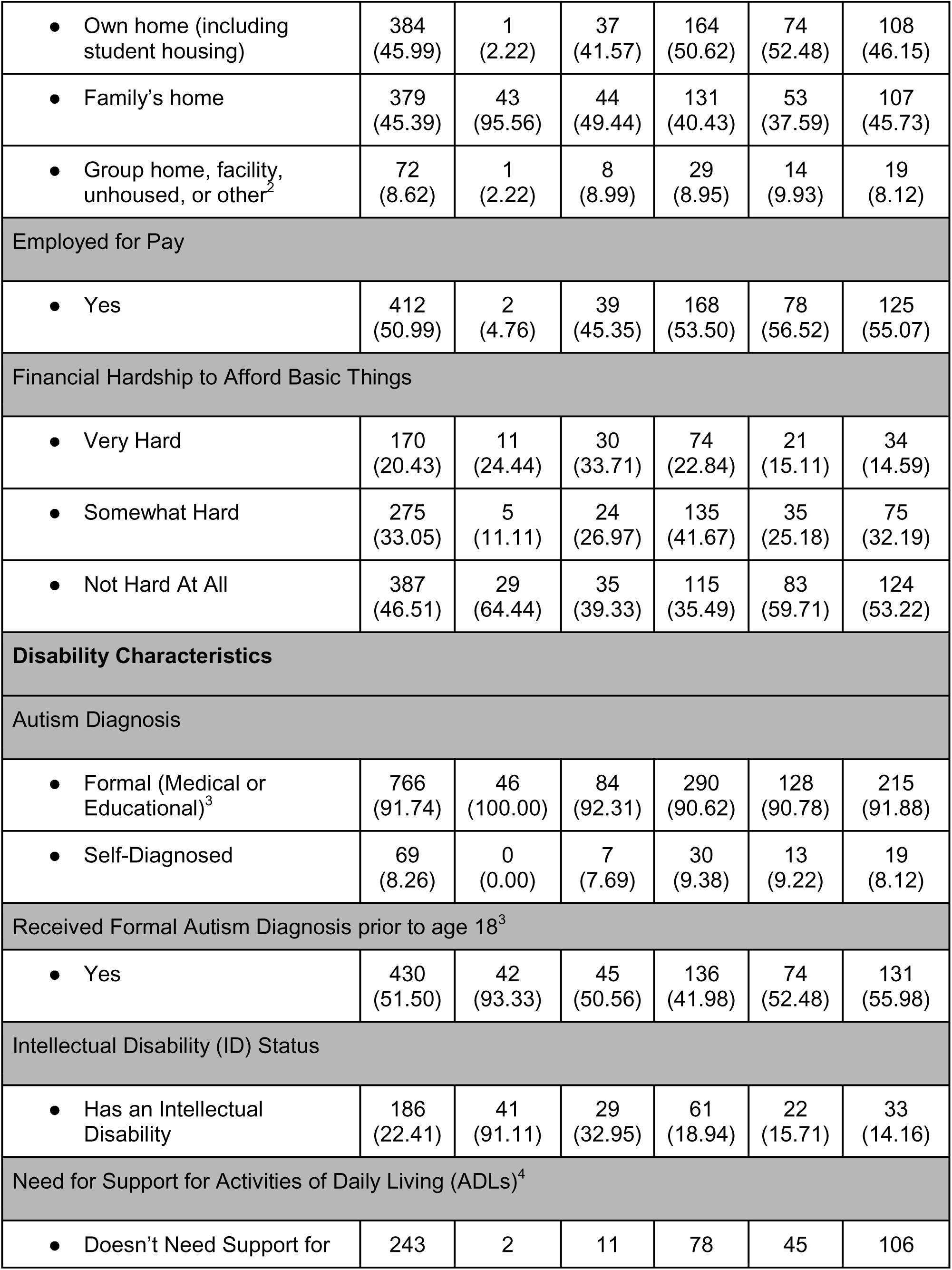
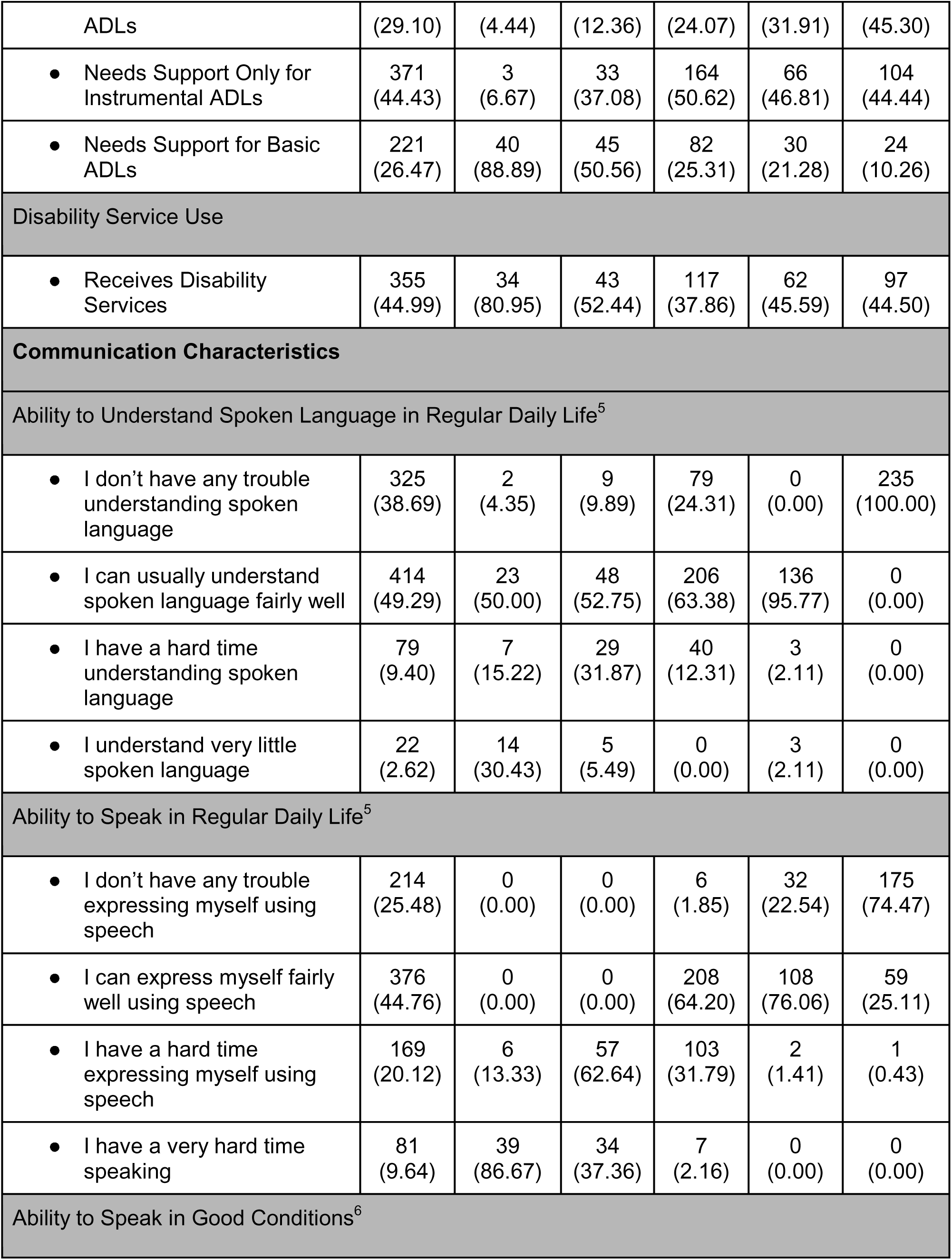
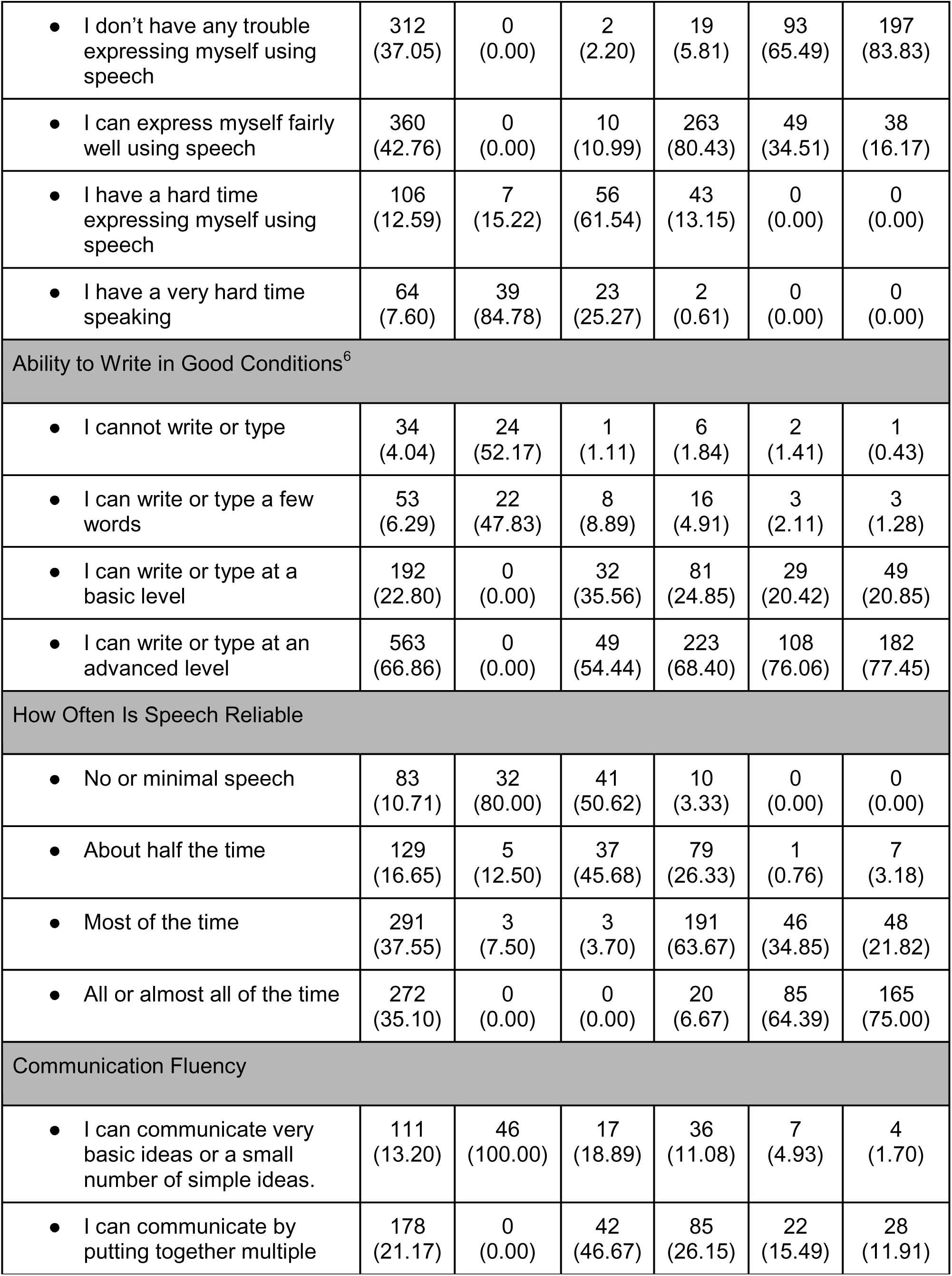
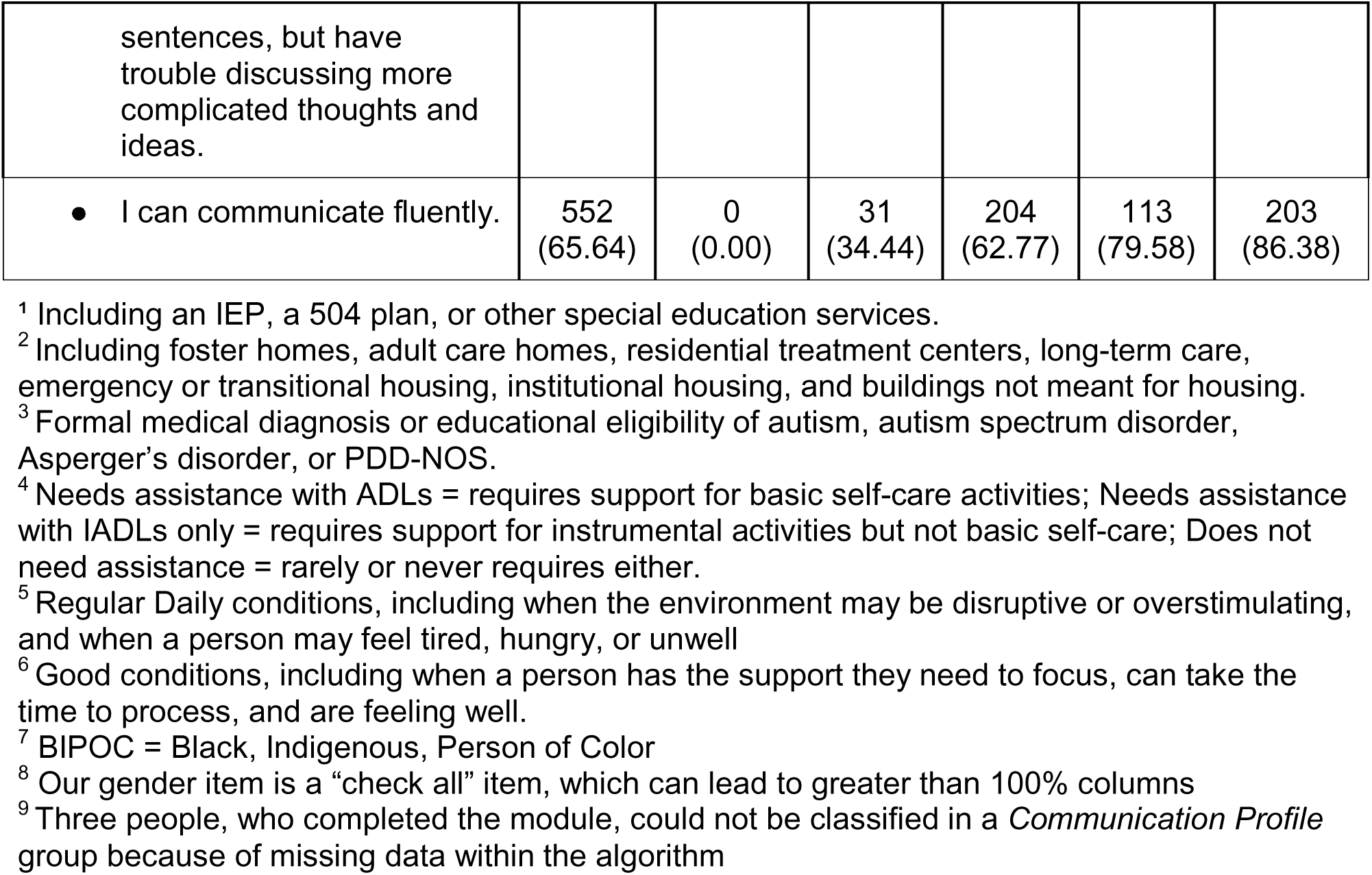
Demographics of the Communication Characteristics Module by Communication Profile.

### Psychometric Evaluation of the Communication Characteristics Module

#### Completion Feasibility and Missing Data

Question completion rates were high across the module (Table 3). For eight universally administered communication questions, completion ranged from 99.5% to 99.8%. The conditionally administered Alternative Forms of Communication item (Question eight) had a completion rate of 97.4% among eligible participants (490/503). The only item with notable missingness was the item on Speech Reliability (Question seven), with 8.18% missing (69/844).

Missingness on Question seven did not differ significantly by survey wave (χ^2^(3) = 5.59, *p* = 0.133), respondent version (χ^2^(2) = 2.00, *p* = 0.367), *Communication Profile* (χ^2^(4) = 7.09,*p* = 0.131), speaking ability in good conditions (χ^2^(3) = 3.57, *p* = 0.312), or speaking ability in daily life (χ^2^(3) = 2.31, *p* = 0.511). Similar findings were observed when analyses were restricted to baseline respondents, suggesting that nonresponse was not systematically associated with participant characteristics.

#### Alternative Forms of Communication

Among the 503 respondents eligible for the Alternative Forms of Communication item, most endorsed at least one alternative communication strategy. Approximately half (52.5%) selected one method and 45.0% selected two or more. Only 13 participants (2.6%) did not endorse any listed alternatives. A frequency table of endorsed alternative forms of communication is provided in Supplement B.

#### Qualitative Review of Alternative Communication Write-In Responses

Additionally, sixty-seven respondents provided write-in responses to the “Other” category. Direct-report respondents frequently described compensatory strategies, including speech-generating devices and text-to-speech applications, writing, digital communication tools, gestures, trusted communication partners, repetition, and communication withdrawal or avoidance. Many emphasized the internal experience of communication difficulty, frustration, inability to communicate, and reliance on compensatory strategies during communication breakdowns.

Caregiver-report respondents more commonly described observable behaviors and partner-mediated communication, including interpretation of behaviors, facial expressions, routines, gestures, limited verbal approximations, and communication supports such as choice boards or AAC devices.

Few write-in responses described communication methods not already represented in the response options. Most could be categorized within existing AAC, partner-assisted communication, gesture interpretation, or writing-based categories, suggesting that the item captured most alternative communication strategies used by participants.

#### Digital Communication Modalities

Digital communication was widely used throughout the sample (Supplement C). Text messaging was the most frequently endorsed modality (85.78%), followed by email (74.53%), telephone communication (63.86%), and videoconferencing (61.02%). Only 9.12% reported that none of the listed digital modalities were reliable.

#### Construct Validity

##### Relationship Between Module Items

Relationship testing demonstrated strong associations between related module items, with all hypothesized associations statistically significant (*p* < .001). The analyses identified significant associations between understanding speech and literacy, matched items assessing communication in good conditions versus daily life, reading versus writing ability, and speech reliability versus overall communication fluency. The direction and pattern of these associations aligned with the prespecified hypotheses (Table 2).

**Table 2.** Relationship Between Communication Characteristics Module Items.

| <u>Hypothesis</u> | <u>Prediction</u> | <u>Strength</u> | <u>Result</u> |
| --- | --- | --- | --- |
| Q1: Ability to understand speech in good conditions<sup>1</sup><br>vs.<br>Q2: Ability to understand speech in regular daily life<sup>2</sup> | large | large | [$\chi^2(9) = 954.96$,<br>$p < .001$, Cramér's $V = 0.62$] |
| Q1: Ability to understand speech in good conditions<sup>1</sup><br>vs.<br>Q5: Ability to read in good conditions<sup>1</sup> | large | large | [$\chi^2(9) = 244.95$,<br>$p < .001$, Cramér's $V = 0.31$] |
| Q3: Ability to speak in good conditions<sup>1</sup><br>vs.<br>Q4: Ability to speak in regular daily life<sup>2</sup> | large | large | [$\chi^2(9) = 1066.65$,<br>$p < .001$, Cramér's $V = 0.65$] |
| Q3: Ability to speak in good conditions<sup>1</sup><br>vs.<br>Q10: Communication fluency in good conditions in best modality<sup>1,3</sup> | large | large | [$\chi^2(9) = 415.03$,<br>$p < .001$, Cramér's $V = 0.41$] |
| Q5: Ability to read in good conditions<sup>1</sup><br>vs.<br>Q6: Ability to write in good conditions<sup>1</sup> | large | large | [$\chi^2(9) = 1086.05$,<br>$p < .001$, Cramér's $V = 0.66$] |
| Q7: Frequency speech is a reliable form of communication<br>vs.<br>Q10: Communication fluency in good conditions in best modality<sup>1,3</sup> | large | large | [$\chi^2(12) = 197.57$,<br>$p < .001$, Cramér's $V = 0.29$] |

##### Known-Groups Validity

As hypothesized, response distributions differed significantly depending on whether responses were direct self-report, with or without support, or caregiver report for all primary communication items (Questions one through seven and 10, all *p* < .001). Caregiver reports consistently indicated greater communication difficulties than direct self-reports, while supported self-reports generally fell between these groups. Effect sizes ranged from moderate to large. The strongest associations were observed for writing ability (Cramér’s *V* = .512), reading ability (*V*= .486), overall communication fluency (*V* = .457), and use of speech in good conditions (*V*= .411). Moderate effects were observed for use of speech in daily life (*V* = .328), speech reliability (*V*= .276), understanding speech in good conditions (*V*= .252), and understanding speech in daily life (*V*= .238).

These findings support the construct validity of the Communication Characteristics module and demonstrate that item responses differentiate groups expected to vary in communication support needs.

#### Test-Retest Reliability

We evaluated item-level test-retest reliability using weighted Cohen’s kappa coefficients based on the 6-month survey and 2-week retest. Reliability was substantial for all ordinal items (Table 3). Weighted kappas ranged from .63 to .75 for receptive language, expressive communication, speech reliability, and overall communication fluency. Literacy items demonstrated the strongest reliability, with reading ability showing almost perfect agreement (κ = .89) and writing ability demonstrating excellent agreement (κ = .84). These findings indicate acceptable temporal stability across all module items.

**Table 3.** Item-Level Completion Rates and Test-Retest Reliability for the Communication Characteristics Module.

| **Question** | **N Eligible** | **% Missing** | **Unweighted $K^*$** | **Weighted $K^*$** | **% Agreement** | **Interpretation** |
| --- | --- | --- | --- | --- | --- | --- |
| 1 - Understanding Speech in Good Conditions | 844 | 0.24 | 0.598 | 0.648 | 91.8% | Substantial |
| 2 - Understanding Speech in Daily Life | 844 | 0.47 | 0.582 | 0.627 | 90.9% | Substantial |
| 3 - Using Speech in Good Conditions | 844 | 0.24 | 0.580 | 0.711 | 91.1% | Substantial |
| 4 - Using Speech in Daily Life | 844 | 0.47 | 0.599 | 0.709 | 90.6% | Substantial |
| 5 – Reading in Good Conditions | 844 | 0.36 | 0.833 | 0.891 | 97.1% | Almost perfect |
| 6 – Writing in Good Conditions | 844 | 0.24 | 0.753 | 0.843 | 95.8% | Almost perfect |
| 7 – Speech as a Reliable Form of Communication | 844 | 8.18 | 0.583 | 0.680 | 88.6% | Substantial |
| 8 – Alternative Form of Communication<sup>1</sup> | 503 | 2.58 | NA | NA | NA | NA |
| 9 – Use of Digital Communication<sup>1</sup> | 844 | 0.24 | NA | NA | NA | NA |
| 10 – Overall Communication in Good Conditions in Preferred Modality | 844 | 0.36 | 0.682 | 0.749 | 92.3% | Substantial |

*\*Note: $K$ = Cohen's kappa. Weighted kappas were calculated using linear weights. Interpretation categories follow commonly accepted benchmarks for agreement.*
*<sup>1</sup>Items 8 (Alternative Form of Communication) and 9 (Use of Digital Communication) were check-all-that-apply items and were not evaluated using kappa statistics.*

### Psychometric Evaluation of the Communication Profile Variable

#### Missing Profiles

The *Communication Profile* algorithm was successfully calculated for 841 of 844 baseline participants (99.6%) (Table 1). Three participants could not be assigned a category because of missing responses required for profile determination. One lacked sufficient information across items assessing speech use in good conditions, speech use in daily life, and speech reliability, while two were missing information about understanding speech in daily life.

#### Test-Retest Reliability

The derived *Communication Profile* variable demonstrated substantial test-retest agreement (κ = .69) using linear weighted kappa. Transition matrices indicated that disagreements most commonly occurred between adjacent categories. Participants classified as “Minimal Speakers without robust alternative communication” and “Speakers without difficulty producing or understanding speech” showed the greatest stability, whereas transitions were more common among intermediate profiles. These findings suggest that the *Communication Profile* captures relatively stable communication characteristics.

#### Construct Validity

We evaluated construct validity of the *Communication Profile* variable through a series of *a priori* hypothesis tests developed during the community-based profile development process. All hypotheses were supported (Table 4). Minimal Speakers with and without a robust alternative communication differed significantly in the expected direction in respondent version, reading ability, and intellectual disability status, with effect sizes ranging from medium to very large (Cramér’s *V* = .40-.81). However, while receptive language ability was associated with *Communication Profile*, the two minimal speaker profiles differed in their distributions, with neither profile consistently demonstrating greater receptive language ability across response categories.

**Table 4.** A priori hypothesis testing for Communication Profile variable.

|  |  |
| --- | --- |
| **Communication Profiles Tested:** |  |
| Minimal Speakers without robust alternative communication (Communication Profile = 0)<br>vs.<br>Minimal Speakers with robust alternative communication (Communication Profile = 1) |  |
| **Hypothesis:** | **Result:** |
| Profile 0 will be more frequently represented by caregiver reports, whereas Profile 1 will be more frequently represented by direct reports. | $[\chi^2(2) = 55.14, p < .001, \text{Cramér's } V = .63]$ |
| Profile 1 will demonstrate greater reading ability than Profile 0. | $[\chi^2(3) = 89.96, p < .001, \text{Cramér's } V = .81]$ |
| Profile 0 will include a greater proportion of participants who disclose a co-occurring intellectual disability. | $[\chi^2(1) = 40.39, p < .001, \text{Cramér's } V = .55]$ |
| Profile 0 and Profile 1 will be associated with receptive language ability. | $[\chi^2(3) = 21.51, p < .001, \text{Cramér's } V = .40]$ |
| **Communication Profiles Tested:** |  |
| Speakers with difficulty producing speech (Communication Profile = 2)<br>vs.<br>Speakers without difficulty producing but with difficulty understanding speech (Communication Profile = 3)<br>vs.<br>Speakers without difficulty producing or understanding speech (Communication Profile = 4) |  |
| **Hypothesis:** | **Result:** |
| Communication Profiles 2, 3, and 4 will differ in their reported communication ability under good conditions. | $[\chi^2(6) = 56.99, p < .001, \text{Cramér's } V = .20]$ |
| Participants in Profiles 2, 3, and 4 will differ in reports that communication partners do not give them enough time to communicate successfully, with Profile 2 expected to report this barrier most frequently and Profile 4 least frequently. | $[\chi^2(8) = 71.348, p < .001, \text{Cramér's } V = .23]$ |
| **Communication Profiles Tested:** |  |
| Minimal Speakers (Communication Profiles = 0 and 1)<br>vs.<br>Speakers with difficulty producing speech (Communication Profile = 2)<br>vs.<br>Speakers without difficulty producing speech (Communication Profile = 3 and 4) |  |
| **Hypothesis:** | **Result:** |
| Communication Profile groupings will be associated with level of support needed for activities of daily living, with greater support needs expected among minimal speakers (Profiles 0 and 1) than among speakers with difficulty producing speech (Profile 2) and speakers without difficulty producing speech (Profiles 3 and 4) | $[\chi^2(4) = 138.20, p < .001, \text{Cramér's } V = .29]$ |
| **Communication Profiles Tested:** |  |
| All Communication Profiles Levels |  |
| **Hypothesis:** | **Result:** |
| The amount of support needed to communicate with healthcare providers will differ across Communication Profiles, with participants with greater communication support needs requiring more assistance from supporters during healthcare visits. | $[\chi^2(8) = 206.52, p < .001, \text{Cramér's } V = .36]$ |
| **Communication Profiles Test:** |  |
| Minimal Speakers (Communication Profiles = 0 and 1)<br>vs.<br>Speakers (Communication Profiles = 2, 3, and 4) |  |
| **Hypothesis:** | **Result:** |
| Minimal Speakers (Profiles 0 and 1) will report greater overall barriers to communication than speakers in Profiles 2 through 4. | $[t(839) = 9.65, p < .001, \text{Cohen's } d = 1.18]$ |

The three speaking *Communication Profile* groups also differed significantly in overall communication fluency, communication barriers related to insufficient time to communicate, support needs for activities of daily living, and support required during healthcare visits. Associations with communication fluency and communication barriers were modest (Cramér’s *V* = .20 - .23), while differences in functional support needs and healthcare communication support were moderate (Cramér’s *V* = .29-.36), consistent with hypothesized gradients in communication-related support needs.

Finally, minimally speaking participants (Profiles 0-1) reported substantially greater overall communication barriers than speaking participants (Profiles 2-4). Mean barriers-to- communication scores were nearly twice as high among minimally speaking participants (*M* = 123.80, *SD* = 67.34) as speakers (*M* = 65.93, *SD* = 44.78), representing a large effect (Cohen’s *d* = 1.18). A Mann-Whitney U test yielded consistent results (*z* = 10.47, *p* < .001).

Taken together, these findings provide strong support for the construct validity of the *Communication Profile* variable across measures of communication ability, literacy, intellectual disability status, functional support needs, healthcare communication support, and overall communication barriers.

## Discussion

This study provides evidence that the AASPIRE Communication Characteristics module is a reliable and valid instrument for characterizing communication heterogeneity among autistic adults. The module was intentionally developed as a multidimensional demographic characterization instrument rather than a unidimensional scale, encompassing receptive and expressive communication, literacy, speech reliability, alternative communication modalities, digital communication, and overall communication fluency. Meaningful variation across these domains, combined with expected relationships between related constructs, supports this approach.

A major contribution from our work is the development and validation of the *Communication Profile* variable. Our algorithm classified participants into five conceptually meaningful groups that differed in expected ways across reading ability, intellectual disability status, support needs, healthcare communication support, communication barriers, and respondent type. These findings suggest that our profile variable captures meaningful demographic variation in communication experiences beyond simple distinctions based on the presence or absence of speech.

Historically, autism research has often characterized communication using binary classifications such as speaking versus nonspeaking (or, more crudely, verbal versus nonverbal). These distinctions obscure heterogeneity within both groups. Individuals who use spoken language vary in speech reliability, ability to communicate complex ideas, and ability to understand spoken language. Similarly, minimally speaking individuals vary in their ability to communicate through writing, AAC, or other modalities. Our *Communication Profile* addresses these limitations by incorporating multiple dimensions of communication and generating categories more reflective of lived communication experiences.

The findings among minimally speaking participants are particularly important. Participants with robust alternative communication differed substantially from those without robust alternative communication in literacy, intellectual disability status, receptive language, and respondent type. These findings demonstrate that minimally speaking autistic adults are not a homogeneous population and that communication ability cannot be inferred from speech alone. Whether these differences reflect developmental pathways, access to communication supports, environmental opportunities, or a combination of factors remains unclear (Eicher et al., 2026).

These findings are also relevant to ongoing discussions regarding communication-based subgrouping within autism. The *Lancet* Commission proposed the term “profound autism” to describe autistic individuals with minimal speech, intellectual disability, or substantial support needs, and the Interagency Autism Coordinating Committee proposed a related framework incorporating functional dependence in daily life. Critics have noted that such categories may combine individuals with substantially different communication strengths and challenges (Kripke-Ludwig, 2023). Our findings support these concerns by identifying meaningful distinctions within minimally speaking participants and among individuals who use speech but experience significant communication challenges. Communication functioning may therefore be better represented through multidimensional characterization than broad labels that merge diverse populations.

Importantly, communication-related support needs were not limited to minimally speaking autistic adults. Participants who used speech but reported difficulty expressing themselves or understanding spoken language experienced elevated communication barriers and often required additional support. This has implications for healthcare, education, employment, and disability services, where eligibility and support decisions may rely on indicators such as speech status solely. Communication challenges can persist even among individuals who appear verbally fluent.

The CBPR process represents another important strength of our work. Rather than relying only on clinician-defined communication constructs, autistic adults with diverse communication experiences helped identify meaningful domains, develop items, and refine profile categories. This contributed to a measure that emphasizes communication experiences and participation rather than deficits alone and aligns with calls for autism research tools developed in partnership with autistic communities.

More broadly, our work illustrates the value of moving beyond single-variable characterizations of autism. Similar approaches could be applied to other domains of heterogeneity, including sensory processing, motor planning, emotional regulation, executive functioning, and cognition. Such approaches may provide more precise ways to understand variation and identify supports, accommodations, and interventions for specific groups of autistic people.

### Limitations

Several limitations should be considered. First, although the module captures multiple dimensions of communication, it does not comprehensively assess all aspects relevant to autistic adults. In particular, it does not include questions about language pragmatics, nonverbal communication such as facial expressions and prosody, or gesture production or interpretation, nor did it identify speech which is echolalic, scripted, or content limited. Future iterations could incorporate these domains.

Second, the *Communication Profile* variable was designed as a practical stratification tool for research, clinical care, and services rather than as a comprehensive model of communication functioning. To maintain interpretability, the algorithm collapsed related communication constructs into five broad categories, including aspects of speech reliability and expressive communication across conditions. Thus, the profile reflects the effectiveness of supports and communication conditions currently available to an individual, rather than the supports they may need or have not yet accessed; differences in access to AAC, technology, or communication partners may therefore influence profile categorization. This may obscure distinctions between individuals whose speech is highly complex but inconsistent and those whose speech is consistently limited. Future studies should examine whether additional profile levels or alternative algorithms better capture these distinctions.

The Speech Reliability item also demonstrated higher missingness than other module items. Approximately 8-10% of participants across study waves did not respond to the item, although missingness was not systematically associated with respondent version, *Communication Profile*, or expressive communication level. Because this item contributes to the profile algorithm, future research should investigate whether wording, response options, or conceptual framing contributed to nonresponse.

The qualitative review of alternative communication write-ins identified additional areas for refinement. Although predefined categories captured most reported communication supports, respondents also described withdrawal and avoidance, emotional or behavioral communication, creative expression, and digital supports such as artificial intelligence or text-based applications. The review also suggests differences between self- and caregiver-report perspectives: caregivers more often described observable behaviors and partner-mediated interpretation, whereas direct-report respondents more often described communication breakdown, exhaustion, compensation, and social consequences. Continued inclusion of both perspectives may therefore be important in communication research.

Finally, minimally speaking autistic adults and caregiver-report participants remained underrepresented relative to population estimates. Most participants completed direct self-report and reported relatively strong literacy skills, which may limit generalizability to autistic adults with greater communication or cognitive support needs. Future studies should prioritize targeted recruitment of minimally speaking autistic adults, AAC users, and caregiver-report respondents.

### Conclusion

The Communication Characteristics module and the *Communication Profile* variable provide researchers, clinicians, service providers, and policymakers with practical tools for accounting for communication heterogeneity among autistic people. Their use may improve participant characterization, facilitate more targeted recruitment and subgroup analyses, enhance interpretation of patient-reported outcomes, and support services and interventions aligned with the communication characteristics of autistic adults. By moving beyond simplistic speech-based classifications, these tools provide a more precise framework for understanding communication diversity and advancing research, services, and supports across the autistic community.

## Ethical Considerations

The Institutional Review Board (IRB) at Portland State University (PSU) approved the project and served as the Single IRB (approval # 227674-18); Oregon Health & Science University, Vanderbilt University Medical Center, and San Diego State University waived authorization to the PSU IRB. Involvement by the Multnomah County Developmental Disability Services was subsumed under the PSU IRB.

Consent to participate: Participants or their legally authorized representatives gave written consent online or in-person.

## Availability of data

All data from this study will be available through the National Institutes of Health (NIH) National Data Archive (NDA).

## Acknowledgements

The authors would like to acknowledge the contributions of other members of the AASPIRE Outcomes Project team who made this work possible including Dora M. Raymaker, PhD, Mirah Scharer, BS, Todd Edwards, PhD, Emanuel Frowner, Andrea Joyce, Julia Love, PhD, Katherine McDonald, PhD, Ivanova Smith, Finn Garner, MA, and Reid Caplan, MA

