## Supplement A for "Development of a Communication Characteristics Module for Autistic Adults and the Creation of a *Communication Profile* Variable"

Christina Nicolaidis, MD, MPH and the Outcomes Project Team

### AASPIRE Communication Characteristics Module

*AASPIRE Communication Characteristics Module Overview* 2

**Description** 2

**Development** 2

**Conditions for Use** 2

**References** 2

*AASPIRE Communication Characteristics Module: Direct Report Version* 3

**Module Items** 3

Standard Direct Report Items on Perceived Change and Use of Support to Participate 8

**Hyperlinks** 9

*AASPIRE Communication Characteristics Module: Caregiver Report Version* 11

**Module Items** 11

Standard Caregiver Report Items on Perceived Change and Knowledge Sources 17

**Hyperlinks** 18

Copyright 2025, Academic Autism Spectrum Partnership in Research and Education (AASPIRE)

### AASPIRE Communication Characteristics Module Overview

#### Description

The AASPIRE Communication Characteristics Module collects information about self-reported (or caregiver-reported) receptive, expressive, and overall communication fluency; reading and writing level; and access to and use of alternatives to speech. It uses multiple-choice, check-all, and write-in items.

#### Development

The AASPIRE Outcomes Project is using a community based participatory research (CBPR) approach to 1) create and validate the AASPIRE Measurement Toolkit; and 2) understand what predicts changes in outcomes over time (Nicolaidis et al., 2026). Each instrument in the AASPIRE Measurement Toolkit has self- and caregiver-reported versions. AASPIRE developed and tested the instruments with and for autistic adults with or without intellectual disability, but they could also be used with general populations or other people who may need cognitively accessible survey instruments.

Our community-academic team collaboratively created the AASPIRE Communications Characteristics Module to capture information about communication characteristics that may be important to understand in clinical or research settings.

#### Conditions for Use

The AASPIRE Communication Characteristics Module is available for use with attribution in noncommercial, clinical, and research settings. Permission is required for instrument adaptation, modification, and for-profit use. For permissions inquiries, contact Christina Nicolaidis and AASPIRE at.

Copyright 2025, Academic Autism Spectrum Partnership in Research and Education (AASPIRE)

participatory, real-world, longitudinal study of outcomes in autistic adults. *Autism*, 13623613261476381. https://doi.org/10.1177/13623613261476381

### AASPIRE Communication Characteristics Module: Direct Report Version

#### Module Items

1. In general, how would you describe your ability to **understand spoken language under good conditions**?

a. I don't have any trouble understanding spoken language.

b. I can understand spoken language fairly well.

c. I have a hard time understanding spoken language.

d. I understand very little spoken language (for example, I can only understand simple words or phrases).

2. In general, how would you describe your ability to **understand spoken language in your regular daily life**?

a. I don't have any trouble understanding spoken language.

b. I can understand spoken language fairly well.

c. I have a hard time understanding spoken language.

d. I understand very little spoken language (for example, I can only understand simple words or phrases).

Copyright 2025, Academic Autism Spectrum Partnership in Research and Education (AASPIRE)

3. In general, how would you describe your ability to **speak under good conditions**?

a. I don’t have any trouble expressing myself using speech.

b. I can express myself fairly well using speech.

c. I have a hard time expressing myself using speech.

d. I have a very hard time speaking (for example, I only can use short phrases) or can’t speak at all.

4. In general, how would you describe your ability to **speak in your regular daily life**?

a. I don’t have any trouble expressing myself using speech.

b. I can express myself fairly well using speech.

c. I have a hard time expressing myself using speech.

d. I have a very hard time speaking (for example, I only can use short phrases) or can’t speak at all.

5. How would you describe your ability to **read under good conditions**?

a. I cannot read

b. I can read some words and short sentences

c. I can read at a basic level (for example, I use “Plain Language” or “Easy Read” materials)

d. I can read at an advanced level

Copyright 2025, Academic Autism Spectrum Partnership in Research and Education (AASPIRE)

6. How would you describe your ability to **write or type under good conditions**?

   a. I cannot write or type
   b. I can write or type a few words
   c. I can write or type at a basic level (for example, I can explain simple ideas in writing)
   d. I can write or type at an advanced level

7. How often is speech a reliable form of communication for you?

| Rarely or none of the time | A little of the time | About half the time | Most of the time | All or almost all of the time |
|---|---|---|---|---|
| 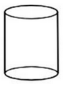 | 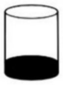 | 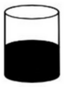 | 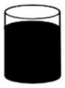 | 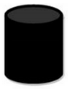 |
| ○ | ○ | ○ | ○ | ○ |

Copyright 2025, Academic Autism Spectrum Partnership in Research and Education (AASPIRE)

*<If "All or almost all of the time" is selected for Question **#7**, skip Question **#8**>*

8. When you **need to use something other than speech to communicate**, what do you do? Please check all that apply

- ❑ I type or write (for example, on paper, a computer, tablet, phone, or other device).
- ❑ I point on a letterboard.
- ❑ I use picture icons (for example, pictures on cards, a computer, tablet, phone, or other device).
- ❑ I use sign language (for example, American Sign Language, ASL).
- ❑ A partner helps me get my thoughts out (for example, they hold a letter board or device, or they provide supports with attention or regulation).
- ❑ Someone tries to interpret my signs, gestures, or behaviors.
- ❑ Someone tries to communicate for me.
- ❑ Other <*write-in*>

9. Which of these things can you **reliably use to communicate**? (check all that apply)

- ❑ Telephone
- ❑ Video-conference (for example, Zoom)
- ❑ Text messaging (for example, on a smart phone)
- ❑ Email
- ❑ None of the above

Copyright 2025, Academic Autism Spectrum
Partnership in Research and Education (AASPIRE)

10. Which of the following statements best describes **your communication, under good conditions, using whatever works best for you** (for example, talking, writing, typing, using sign language, pointing to pictures or letters, or making gestures)?

a. I can only communicate very basic ideas or feelings.
   (For example, I can answer yes or no, indicate I am hungry or in pain, or point to something I want.)

b. I can communicate a small number of simple ideas or feelings.
   (For example, I can use short phrases or sentences or choose among a small number of pictures.)

c. I can communicate by putting together multiple sentences, but I have trouble discussing more complicated thoughts and ideas.

d. I can communicate fluently.
   (For example, I can usually discuss complicated thoughts and ideas.)

Copyright 2025, Academic Autism Spectrum Partnership in Research and Education (AASPIRE)

#### Standard Direct Report Items on Perceived Change and Use of Support to Participate

Overall, in the last six months, did the things this survey asked about...

- ○ Get worse
- ○ Stay about the same
- ○ Get better

Did you get any help while answering the questions in this section of the survey?

- ○ Yes
- ○ No

*<If 'Yes' to get help question>*

What kind of help did you get while answering the questions in this section of the survey? (Check all that apply.)

- ❑ Someone helped me use a computer, smartphone, or other device (for example, they clicked on the answers I chose).
- ❑ Someone read the survey to me.
- ❑ Someone helped me understand what the questions or answers options meant.
- ❑ Someone helped me decide how to answer the questions on the survey.
- ❑ I got some other type of help to answer the questions.
- ❑ Someone answered the questions for me, without my input.

Copyright 2025, Academic Autism Spectrum Partnership in Research and Education (AASPIRE)

#### Hyperlinks

---

Understand spoken language under good conditions

Think about how well you understand speech when you have the support you need to focus, you can take the time to process what is being said, and you are feeling well.

---

Understand spoken language in your regular daily life

This may include times where there are lots of distractions or sensory input, when the conversation is happening quickly, or when you are feeling sick, stressed, or tired.

---

Speak under good conditions

Think about how well you can speak when you have the support you need to focus, you can take the time you need to say what you want, and you are feeling well.

---

Speak in your regular daily life

This may include times where there are lots of distractions or sensory input, when the conversation is happening quickly, or when you are feeling sick, stressed, or tired.

---

Copyright 2025, Academic Autism Spectrum
Partnership in Research and Education (AASPIRE)

#### Read under good conditions

Think about how well you can read when things are set up well for you. For example, when you have the support you need to focus.

#### Write or type under good conditions

Think about how well you can write or type when things are set up well for you. For example, when you have the support, accommodations, or technology you need to handle any physical or motor issues that get in the way.

#### Your communication, under good conditions

Think about how well you can communicate when you have the support you need, you can take the time you need, and you are feeling well.

Copyright 2025, Academic Autism Spectrum Partnership in Research and Education (AASPIRE)

### AASPIRE Communication Characteristics Module: Caregiver Report Version

#### Module Items

1. In general, how would you describe their ability to **understand spoken language, under good conditions**?

a. I don't think they have any trouble understanding spoken language.

b. I think they can usually understand spoken language fairly well.

c. I think they often have a hard time understanding spoken language.

d. I think they understand very little spoken language (for example, they can only understand simple words or phrases).

Copyright 2025, Academic Autism Spectrum Partnership in Research and Education (AASPIRE)

2. In general, how would you describe their ability to **understand spoken language in their regular daily life**?

a. I don’t think they have any trouble understanding spoken language.

b. I think they can usually understand spoken language fairly well.

c. I think they often have a hard time understanding spoken language.

d. I think they understand very little spoken language (for example, they can only understand simple words or phrases).

3. In general, how would you describe their ability to **speak under good conditions**?

a. I don’t think they have any trouble expressing themselves using speech.

b. I think they can express themselves fairly well using speech.

c. I think they have a hard time expressing themselves using speech.

d. I think they have a very hard time speaking (for example, they only can use short phrases) or can’t speak at all.

Copyright 2025, Academic Autism Spectrum Partnership in Research and Education (AASPIRE)

4. In general, how would you describe their ability to **speak in their regular daily life**?

a. I don't think they have any trouble expressing themselves using speech.

b. I think they can express themselves fairly well using speech.

c. I think they have a hard time expressing themselves using speech.

d. I think they have a very hard time speaking (for example, they only can use short phrases) or can't speak at all.

5. How would you describe their ability to **read under good conditions**?

a. I don't think they can read.

b. I think they can read some words and short sentences.

c. I think they can read at a basic level (for example, they use "Plain Language" or "Easy Read" materials).

d. I think they can read at an advanced level.

Copyright 2025, Academic Autism Spectrum Partnership in Research and Education (AASPIRE)

6. How would you describe their ability to **write or type under good conditions**?

a. I do not think they can write or type.

b. I think they can write or type a few words.

c. I think they can write or type at a basic level (for example, they can explain simple ideas in writing)

d. I think they can write or type at an advanced level.

7. How often do you think speech is a reliable form of communication for them?

| Rarely or none of the time | A little of the time | About half the time | Most of the time | All or almost all of the time |
|---|---|---|---|---|
| ○ | ○ | ○ | ○ | ○ |

Copyright 2025, Academic Autism Spectrum Partnership in Research and Education (AASPIRE)

*<If "All or almost all of the time" is selected for Question **#7**, skip Question **#8**>*

8. When they **need to use something other than speech to communicate**, what do they do? Please check all that apply.

- ❑ They type or write (for example, on paper, a computer, tablet, phone, or other device).
- ❑ They point on a letterboard.
- ❑ They use picture icons (for example, pictures on cards, a computer, tablet, phone, or other device).
- ❑ They use sign language (for example, American Sign Language, ASL).
- ❑ A partner helps them get their thoughts out (for example, the partner holds a letter board or device, or provides supports with attention or regulation).
- ❑ Someone tries to interpret their signs, gestures, or behaviors.
- ❑ Someone tries to communicate for them.

9. Which of these things do you think they can **reliably use to communicate**? (check all that apply)

- ❑ Telephone
- ❑ Video-conference (for example, Zoom)
- ❑ Text messaging (for example, on a smart phone)
- ❑ Email
- ❑ None of the above

Copyright 2025, Academic Autism Spectrum Partnership in Research and Education (AASPIRE)

10. Which of the following statements do you think best describes **their communication, under good conditions, using whatever you think works best for them** (for example, talking, writing, typing, using sign language, pointing to pictures or letters, or making gestures)?

a. They can only communicate very basic ideas or feelings. (For example, they can answer yes or no, indicate they are hungry or in pain, or point to something they want).

b. They can communicate a small number of simple ideas or feelings. (For example, they can use short phrases or sentences or choose among a small number of pictures).

c. They can communicate by putting together multiple sentences, but they have trouble discussing more complicated thoughts and ideas.

d. They can communicate fluently. (For example, they can usually discuss complicated thoughts and ideas.)

Copyright 2025, Academic Autism Spectrum Partnership in Research and Education (AASPIRE)

#### Standard Caregiver Report Items on Perceived Change and Knowledge Sources

Overall, in the last six months, did the things this survey asked about...

- ○ Get worse
- ○ Stay about the same
- ○ Get better

How do you know this information?

- ❑ They told me (in whatever mode they communicate).
- ❑ I have observed the action, behavior, or events.
- ❑ I can interpret their behaviors and/or vocalizations.
- ❑ Someone else told me.
- ❑ I feel like I can guess because I have known them for a long time.
- ❑ I feel like I can guess because I spend a lot of time with them.
- ❑ Other *<write-in>*
- ❑ None of the above.

On a scale of 0 to 10, how confident are you in your responses?

*<0 - Not at all Confident to 10 - Totally Confident>*

OPTIONAL: If you are having trouble answering these questions, you may use this space to further clarify or explain your answers. *<write-in>*

Copyright 2025, Academic Autism Spectrum Partnership in Research and Education (AASPIRE)

#### Hyperlinks

---

Understand spoken language under good conditions

Think about how well you think they understand speech when they have the support they need to focus, they can take the time to process what is being said, and they are feeling well.

---

Understand spoken language in your regular daily life

This may include times where there are lots of distractions or sensory input, when the conversation is happening quickly, or when you think they are feeling sick, stressed, or tired.

---

Speak under good conditions

Think about how well they can speak when they have the support they need to focus, they can take the time they need to say what they want, and they are feeling well.

---

Copyright 2025, Academic Autism Spectrum Partnership in Research and Education (AASPIRE)

---

#### Speak in your regular daily life

This may include times where there are lots of distractions or sensory input, when the conversation is happening quickly, or when you think they are feeling sick, stressed, or tired.

---

#### Read under good conditions

Think about how well they can read when things are set up well for them. For example, when they have the support they need to focus.

---

#### Write or type under good conditions

Think about how well they can write or type when things are set up well for them. For example, when they have the support, accommodations, or technology they need to handle any physical or motor issues that get in the way.

---

Copyright 2025, Academic Autism Spectrum Partnership in Research and Education (AASPIRE)

#### Their communication, under good conditions

Think about how well they can communicate when they have the support they need, they can take the time they need, and they are feeling well.

Copyright 2025, Academic Autism Spectrum Partnership in Research and Education (AASPIRE)
