## Supplement B for "Development of a Communication Characteristics Module for Autistic Adults and the Creation of a *Communication Profile* Variable"

| Frequency Table of Alternative Forms of Communication | | | | |
| --- | --- | --- | --- | --- |
| **Alternative Form** | **Checked** | **Unchecked** | **% Checked** | ***n* Eligible** |
| I type or write | 354 | 149 | 70.38% | 503 |
| I point on letterboard | 17 | 486 | 3.38% | 503 |
| I use picture icons | 77 | 426 | 15.31% | 503 |
| I use sign language | 57 | 446 | 11.33% | 503 |
| A partner helps me get my thoughts out | 84 | 419 | 16.70% | 503 |
| Someone tries to interpret my signs, gestures, or behaviors | 133 | 370 | 26.44% | 503 |
| Someone tries to communicate for me | 134 | 369 | 26.64% | 503 |
| Other* | 67 | 436 | 13.32% | 503 |
