## Supplement C for "Development of a Communication Characteristics Module for Autistic Adults and the Creation of a *Communication Profile* Variable"

Reliable Forms of Digital Communication Endorsed by Participants

| **Digital Communication Type** | **Checked** | **Unchecked** | **% Checked** | ***n* Eligible** |
| --- | --- | --- | --- | --- |
| Telephone | 539 | 305 | 63.86 | 844 |
| Video-conferencing (e.g., Zoom) | 515 | 329 | 61.02 | 844 |
| Text Messaging | 724 | 120 | 85.78 | 844 |
| Email | 629 | 215 | 74.53 | 844 |
| None of the above | 77 | 767 | 9.12 | 844 |
