## Supplement D for "Development of a Communication Characteristics Module for Autistic Adults and the Creation of a *Communication Profile* Variable"

**5 Communication Profile Variable Logic**

Step 1. Classify Speakers and Non-Speakers

Using Q3 Q4 and Q7 get an average

- Important: Q7 has more groups than the other two, so the lowest two answers (0 and 1) both get coded as 0
- Important: Make sure all 3 are coded 0-3 and in the right direction - low to high
  - <= 1 minimal speakers = 0 in new *communication profile* variable
  - > 1 and <= 2.33 challenge speakers = 2 in new *communication profile* variable
  - > 2.33 non-challenged speaker = 4 in new *communication profile* variable

Step 2. Classify Minimal Speakers by Robust and Non-Robust

Using Q10 and Q6 create a logic statement

- Important: Make sure both are coded 0-3 (both already in right direction)
- This logic should only be applied to current *communication profile* = 0 people
  - If speech fluency >= 2 or if write_good >=2 then new *communication profile* = 1, else *communication profile* = 0

At the end of Step 2, you should have 4 groups in the new *communication profile* variable

0 = minimal speech without robust alternative form of communication

1 = minimal speech with robust alternative form of communication

2 = difficulty with speech

4 = little or no difficulty with speech

Step 3. Further Classify Speakers by Understanding of Speech in Good Conditions

- Important: This classification should only apply to people in current *communication profile* = 4
  - Of those that are in *communication profile* 4, what are their difficulties with understanding speech?
    - You are essentially creating a “difficulty with understanding spoken speech” bivariate that applies a yes/no logic

Using Q2

- Important: Needs to be coded 0-3 and in the right direction - low to high
  - >= 3 without challenges understanding speech
  - < 3 with challenges understanding speech

Step 4. Apply Understanding Bivariate to *communication profile* 4

At the end of Step 4, you should have all 5 groups in the new *communication profile* variable

**Example Coding for Stata**

*recoding Qs 3, 4, and 7 to zero values and in correct order*

gen spk_good = comm_spk_good

replace spk_good = 0 if comm_spk_good == 4

replace spk_good = 1 if comm_spk_good == 3

replace spk_good = 2 if comm_spk_good == 2

replace spk_good = 3 if comm_spk_good == 1

gen spk_daily = comm_write_good

replace spk_daily = 0 if comm_write_good == 4

replace spk_daily = 1 if comm_write_good == 3

replace spk_daily = 2 if comm_write_good == 2

replace spk_daily = 3 if comm_write_good == 1

gen spk_reliable0 = comm_reliable

replace spk_reliable0 = 0 if inlist(comm_reliable, 1, 2)

replace spk_reliable0 = 1 if comm_reliable == 3

replace spk_reliable0 = 2 if comm_reliable == 4

replace spk_reliable0 = 3 if comm_reliable == 5

*counting number of missing scores for Qs 3, 4, and 7*

gen miss_spk = missing(spk_good) + missing(spk_daily) + missing(spk_reliable0)

*creating basic mean composite variable for Qs 3, 4, and 7*

gen speech_mean = .

replace speech_mean = (spk_good + spk_daily + spk_reliable0)/3 if miss_spk == 0

*allowing for one missing score in either Q 3, 4, or 7*

replace speech_mean = (spk_good + spk_daily)/2 if miss_spk == 1 & missing(spk_reliable0)

replace speech_mean = (spk_good + spk_reliable0)/2 if miss_spk == 1 & missing(spk_daily)

replace speech_mean = (spk_daily + spk_reliable0)/2 if miss_spk == 1 & missing(spk_good)

*creating first 3 main groups of communication profiles*

gen comm_profile = .

replace comm_profile = 0 if speech_mean <= 1

replace comm_profile = 2 if speech_mean > 1 & speech_mean < 2.33

replace comm_profile = 4 if speech_mean >= 2.33

*creating 0 values for Q10 and Q6*

gen comm_fluency0 = comm_fluency

replace comm_fluency0 = 0 if comm_fluency == 1

replace comm_fluency0 = 1 if comm_fluency == 2

replace comm_fluency0 = 2 if comm_fluency == 3

replace comm_fluency0 = 3 if comm_fluency == 4

gen comm_write_good0 = comm_write_good

replace comm_write_good0 = 0 if comm_write_good == 1

replace comm_write_good0 = 1 if comm_write_good == 2

replace comm_write_good0 = 2 if comm_write_good == 3

replace comm_write_good0 = 3 if comm_write_good == 4

*splitting the minimal speaker groups based on data from Q10 and Q6*

gen alt_com = (comm_fluency0 >= 2 | comm_write_good0 >= 2)

replace comm_profile = 1 if comm_profile == 0 & alt_com == 1

*creating 0 variable from reverse coded Q2 data*

gen understand_daily = comm_understand_spk_dailyr

replace understand_daily = 0 if comm_understand_spk_dailyr == 1

replace understand_daily = 1 if comm_understand_spk_dailyr == 2

replace understand_daily = 2 if comm_understand_spk_dailyr == 3

replace understand_daily = 3 if comm_understand_spk_dailyr == 4

*creating binary variable for Q2 data*

gen understand_challenge = .

replace understand_challenge = 1 if understand_daily < 3

replace understand_challenge = 0 if understand_daily >= 3

*creating final groupings*

replace comm_profile = 3 if comm_profile == 4 & understand_challenge == 1

replace comm_profile = 4 if comm_profile == 4 & understand_challenge == 0
